# Respiratory virus concentrations in human excretions that contribute to wastewater: A systematic review

**DOI:** 10.1101/2023.02.19.23286146

**Authors:** Sarah A. Lowry, Marlene K. Wolfe, Alexandria B. Boehm

## Abstract

Concentrations of nucleic acids from a range of respiratory viruses including human influenza A and B, respiratory syncytial virus (RSV), metapneumovirus, parainfluenza virus, rhinovirus, and seasonal coronaviruses in wastewater solids collected from wastewater treatment plants correlate to clinical data on disease occurrence in the community contributing to the wastewater. Viral nucleic acids enter wastewater from various excretions including stool, urine, mucus, sputum, and saliva deposited in toilets or other drains in buildings. In order to relate the measured concentrations in wastewater at a treatment plant to actual number of infections in a community, concentrations of the viral nucleic acids in these human excretions are needed as inputs to a mass balance model. In this study, we carried out a systematic review and meta-analysis to characterize the concentrations and presence of influenza A and B, respiratory syncytial virus (RSV), metapneumovirus, parainfluenza virus, rhinovirus, and seasonal coronaviruses in stool, urine, mucus, sputum, and saliva. The systematic review protocol can be accessed at https://doi.org/10.17605/OSF.IO/ESVYC. We identified 220 data sets from 50 unique articles that met inclusion criteria and reported information on viral concentrations and presence in these excretions. Data were unevenly distributed across virus type (with the most available for influenza) and excretion type (with the most available for respiratory excretions). The majority of data sets only reported the presence or absence of the virus in an excretion in a cross-sectional study design. There is a need for more concentration data, including longitudinal data, across all respiratory virus and excretion types. Such data would allow quantitatively linking virus wastewater concentrations to numbers of infected individuals.

## Introduction

Respiratory viruses are responsible for millions of infections around the world each year. In 2019, acute lower respiratory infections were the leading cause of death globally in children under five years old (1). Several key viruses are responsible for the majority of respiratory viral infections: human rhinovirus, human parainfluenza viruses 1, 2, 3, and 4, influenza viruses A and B, respiratory syncytial virus (RSV), metapneumovirus, human coronaviruses 229E, HKU1, OC43, and NL63, adenovirus, human bocavirus, and non-rhinovirus enterovirus (2). For example, human rhinoviruses are the most significant causes of the common cold, influenza A and B viruses cause annual epidemics, and RSV and metapneumovirus are estimated to have infected almost all children by the time they reach the age of five (3). Additionally, the recent SARS-CoV-2 pandemic has underscored the public health threat that respiratory viruses pose.

The detection of SARS-CoV-2 RNA in wastewater has led to the rapid growth of wastewater-based epidemiology (WBE) as a tool to aid public health officials in identifying population-level trends throughout the SARS-CoV-2 pandemic. More recently, wastewater surveillance of RSV (4) and influenza (5) has been shown to be strongly correlated with the clinical incidence of these diseases. Additionally, rhinovirus (6), parainfluenza viruses, metapneumovirus, and seasonal coronaviruses 229E, HKU1, OC43, and NL63 have all been detected in wastewater where they are correlated to clinical measures of disease occurrence (7), indicating that wastewater surveillance of respiratory viruses may be a viable tool for public health officials to implement alongside conventional surveillance methods.

While WBE is useful in identifying community-level trends in infections, there is a lack of information on how to translate viral nucleic-acid concentrations measured in wastewater directly to aggregated case numbers. The ability to make this translation would significantly increase the power of WBE by enabling one to estimate disease occurrence directly from wastewater. Many factors and variables influence the potential translation from viral nucleic acid quantities in wastewater to the number of cases (5,8), but perhaps one of the variables with the most uncertainty is the virus concentration in human excretions that contribute to wastewater. These excretions include not only stool and urine but also mucus, saliva, and sputum. Thus, the aim of this systematic review is to characterize respiratory virus concentrations in excretions that contribute to wastewater and identify critical knowledge gaps for further research. Here, we present a characterization of respiratory virus concentration and presence across excretion types from studies examining subjects with respiratory virus infections.

## Methods

### Systematic Review

The systematic review followed PRISMA guidelines (9). This review aimed to gather and synthesize the existing literature on shedding patterns and concentrations of respiratory viruses in the various excretions that contribute to wastewater. The respiratory viruses included in this review were human rhinovirus, human parainfluenza viruses 1, 2, 3, and 4, influenza viruses A and B, respiratory syncytial virus (RSV), metapneumovirus, and human coronaviruses 229E, HKU1, OC43, and NL63. The excretions considered in the review were mucus, saliva, sputum, urine, and stool. The primary goal of the review was to compile concentrations, in units of virus or viral genomes or viral genes per mass or volume of excretion, of respiratory viruses measured in stool, urine, sputum, mucus, and saliva.

The review protocol for this systematic review can be found at https://doi.org/10.17605/OSF.IO/ESVYC (10). Searches were conducted for each respiratory virus between June and August 2022 in the following three databases: Web of Science (search field = topic), PubMed (search field = title/abstract), and Scopus (search field = title/abstract/keywords). The search string consisted of two fields. The first field was the name of the respiratory virus and any common variations or abbreviations of that name, and the second field contained the list of excretions of interest. The first field changed for each respiratory virus and is shown in Table 1. The second field remained constant for each respiratory virus and was as follows: (urine OR feces OR faeces OR fecal OR stool OR sputum OR mucus OR saliva). Once searches were conducted, records were uploaded into Covidence, a web-based software platform made for systematic and literature reviews (11). In Covidence, records were deduplicated, screened, and extracted for data analysis. The inclusion criteria were as follows: each record had to (1) be published in English, (2) be published in a peer-reviewed journal, (3) contain primary data (i.e. not reviews), (4) contain extractable shedding data, where “shedding data” is defined as measured concentrations or presences of viruses or their genetic material in urine, stool, sputum (including expectorated and induced sputum), mucus (including nasal and nasopharyngeal aspirates), or saliva; data had to be from direct excretion measurements and not from areas of the body that generate excretions (e.g. swabs and lavages) and concentrations had to be presented in externally valid units (some measure of virus numbers per volume or mass of excretion and not per mass or volume of nucleic-acid or nucleic acid extract, viral transport media, or other media), (5) contain shedding data from humans, and (6) contain shedding data from subjects infected with the virus of interest or with respiratory and/or gastrointestinal symptoms and not related to a chronic health condition (i.e. COPD, cystic fibrosis). Inclusion criteria were chosen to ensure only credible and original data relevant to the aim of this systematic review were eligible for inclusion. First articles were screened using their title and abstract and if deemed to potentially fit the inclusion criteria, they were subject to full text review. Articles that fit the inclusion criteria passed full text review and were included in the systematic review.

**Table 1.** Search terms for each respiratory virus and the date searched for each respiratory virus. Date is in MM/DD/YY format.

| Respiratory Virus | Search terms | Date Searched |
| --- | --- | --- |
| Rhinovirus | rhinovirus* OR HRV* | 06/22/22 |
| Human Parainfluenza Viruses 1, 2, 3, and 4 | parainfluenza* OR HPIV* OR PIV | 07/13/22 |
| Influenza A and B | "influenza A" OR "influenzavirus A" OR "type A influenza" OR IAV OR "A flu" OR "flu A" OR "influenza B" OR "influenzavirus B" OR "type B influenza" OR IBV OR "B flu" OR "flu B" | 07/20/22 |
| Respiratory Syncytial Virus | RSV OR "respiratory syncytial virus" OR "human orthopneumovirus" | 07/28/22 |
| Human Metapneumovirus | metapneumovirus* OR "human metapneumovirus" OR hMPV OR HMPV OR MPV | 08/25/22 |
| Human Coronaviruses 229E, HKU1, NL64, and OC43 | "human coronavirus 229E" OR HCoV-229E OR "HCoV 229E" OR 229E OR "human coronavirus NL63" OR HCoV-NL63 OR "HCoV NL63" OR NL63 OR "human coronavirus OC43" OR HCoV-OC43 OR "HCoV OC43" OR OC43 OR "human coronavirus HKU1" OR HCoV-HKU1 OR "HCoV HKU1" OR HKU1 | 08/29/22 |

The following data were extracted from each paper found to fit the inclusion criteria by an independent author. First, we noted whether data were obtained using a cross-sectional study design (many individuals sampled each one time), or a longitudinal study design (one or more individuals with samples collected at more than one time point post infection onset); we also noted when in some cases, a cross sectional study incidentally sampled one or more individuals more than once. Second, we noted the virus type and subtype if applicable, virus detection method, and excretion type. Third, we extracted from the publication the concentrations of the virus measured in the excretions as well as any information about the time point in the infection of the individual. Fourth, if concentrations were not reported, we noted whether excretions were positive for the virus (positivity rates), as well as any information about the time point in the infection of the individual. A positivity rate was defined in this review as the number of samples that tested positive for the respiratory virus divided by the total number of samples tested (i.e. a positivity rate of 10/20 means out of 20 samples taken, 10 tested positive and 10 tested negative for the presence of the virus). In some cases, information about concentrations or positivity rates were provided as only summary statistics by authors and in those cases, those were extracted and the type of summary statistic was noted. If a study reported shedding data only in a graphical format, WebPlotDigitizer (12) was used for data extraction.

### Meta-analysis

Data sets from longitudinal and cross sectional studies were reported separately for both positivity rates and concentrations. For each data set, the population studied was categorized as (1) subjects with confirmed infections of the respiratory virus in question, or (2) subjects without confirmed infections but with respiratory and/or gastrointestinal symptoms. A confirmed infection was defined as the positive detection of the virus in at least one clinical sample out of at least two paired samples (for example, if considering positivity rates in stool in a study that examined stool and nasal swabs, confirmed infections were individuals with positive stool samples, nasal swabs, or both).

Data sets from studies with subjects without confirmed infections but with respiratory and/or gastrointestinal symptoms were used to supplement the primary analysis that focused on subjects with confirmed infections. These data sets were supplementary because subjects without confirmed infections but with symptoms may not have been infected with a viral pathogen rather than infected with a viral pathogen but not shedding that pathogen in excretions, and thus were considered separately than data sets of subjects with confirmed infections. These data sets of subjects without confirmed infections were used to provide evidence that viral shedding is possible in excretions that were not examined by data sets of subjects with confirmed infections.

Reported concentrations of viruses were represented graphically for comparison among different data sets and types of excretions. Concentrations from cross-sectional studies were graphed separately from concentrations from longitudinal studies. Positivity rates from longitudinal studies were reported individually and separately from positivity rates from cross-sectional studies. Positivity rates from cross-sectional studies for each excretion type were combined across data sets using a weighted average. The weighted average was calculated in the statistical computing program R (13) using the following formula: 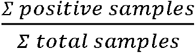 where the numerator represents the sum of positive samples across relevant data sets and the denominator represents the sum of all samples tested across relevant data sets. We tested whether the excretion positivity rates were the same within viruses using either a chi-square test for non-sparse data (i.e. 20% or less of values in the table were less than 5) or a Fisher’s exact test for sparse data (i.e. more than 20% of values in the table were less than 5). We completed the statistics using RStudio (version 2022.07.1) using R (version 4.1.2) (13), as pre-specified in the protocol registration. While some data points were correlated as some excretion samples came from the same subject (e.g. when a study collected paired samples), we were unable to account for correlation in comparing samples across all excretion types. We conducted 6 hypothesis tests and therefore to achieve alpha = 0.05 in the hypothesis tests, we used a p value of 0.008 (0.05/6) to adjust for multiple comparisons (Bonferroni correction). All data used in this paper are available publicly through the Stanford Data Repository (https://doi.org/10.25740/vj779wy5347).

## Results

### Systematic review results

The search process identified a total of 220 data sets from 50 unique articles published in peer-reviewed journals. Figure 1 shows the search process for each respiratory virus. A data set is defined as concentration or positivity rate data for one virus type (e.g. a study reporting both Influenza A and Influenza B data in an excretion would have two data sets) in a specific excretion. If an article provided data for multiple excretions, then data from each excretion made up a unique data set. Many articles included data on multiple respiratory viruses.

**Figure 1.**
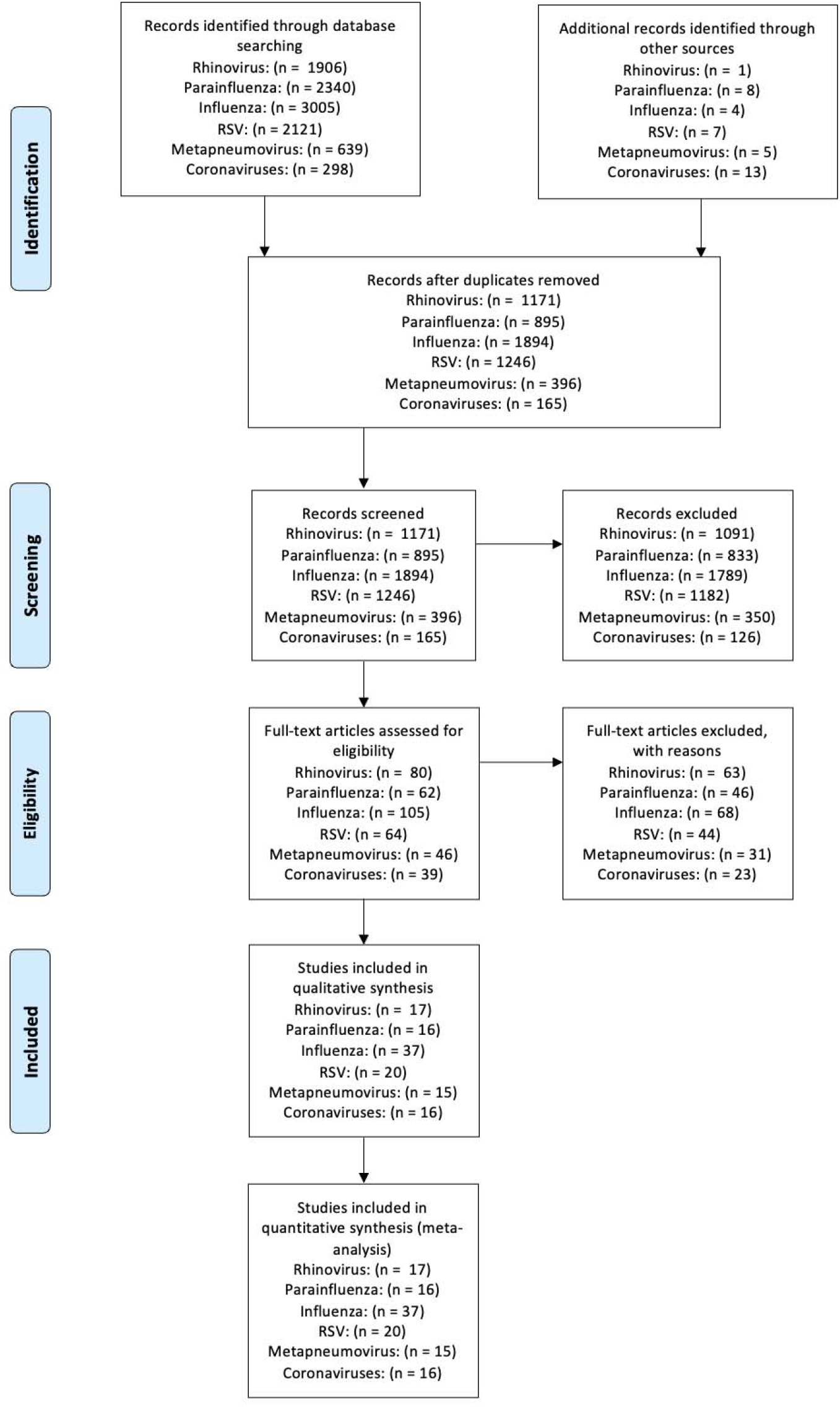
Modified PRISMA flow diagram showing the results of the systematic review search process.

#### Rhinovirus

We identified 17 papers (14–30) with data on rhinovirus in excretions of patients; these papers contained 20 rhinovirus data sets. Of the 20 data sets, one reported longitudinal positivity rates, three reported concentrations from cross-sectional studies, and the remaining 16 data sets reported positivity rates from cross-sectional studies that included between three (29) and 312 (26) subjects with confirmed infections. Of the 20 data sets, 15 measured the virus using RT-PCR-based methods (14,15,17,20–30) while the remaining five used cultivation methods (16,18,19).

The longitudinal data set reported rhinovirus positivity rates in mucus in 26 patients infected with rhinovirus infections over eight weeks (21). The study indicated 26/26 (100%) detects on Day 0, 13/26 (50%) detects at two weeks follow up, 1/26 (4%) detects at five weeks follow up, and 0/26 detects at eight weeks follow up.

Only three data sets reported rhinovirus concentrations in excretions; one data set reports only one datapoint and the remaining two report only summary statistics (Table 2). Data sets reported a median concentration of on the order of 10^3^ genomes per mg stool from 11 subjects, 100 TCID_50_ (median tissue culture infectious dose) per g stool from one subject, and a geometric mean of on the order of 10 TCID_50_ per ml saliva from 7 subjects. There are too few data to complete a statistics test to compare concentrations across excretions, particularly since most of the studies only reported summary statistics.

**Table 2.** Concentrations of rhinovirus reported in excretions. IQR is interquartile range.

| Excretion Type | Data Format | Concentration | Units | Number of subjects | Publication |
| --- | --- | --- | --- | --- | --- |
| Stool | Median (IQR) | 4124 (9117) | genomes/mg | 11 | Bergallo et al. 2019 (14) |
| Stool | Single sample | 100 | TCID<sub>50</sub>/g | 1 | Cate et al. 1967 (16) |
| Saliva | Geometric mean | 15.85 | TCID<sub>50</sub>/mL | 7 | Gwaltney et al. 1978 (18) |

The weighted average percentage of positive samples identified in each excretion type of subjects with confirmed rhinovirus infections is provided in Figure 2, as calculated from the 16 positivity rate data sets from cross-sectional studies. The highest positivity rate was found in mucus samples, while the lowest was found in stool. No studies of subjects with confirmed infection reported measurements of rhinovirus in urine. Fisher’s exact test resulted in a p-value of less than 0.001 rejecting the null hypothesis that positivity rates across excretions are the same.

**Figure 2.**
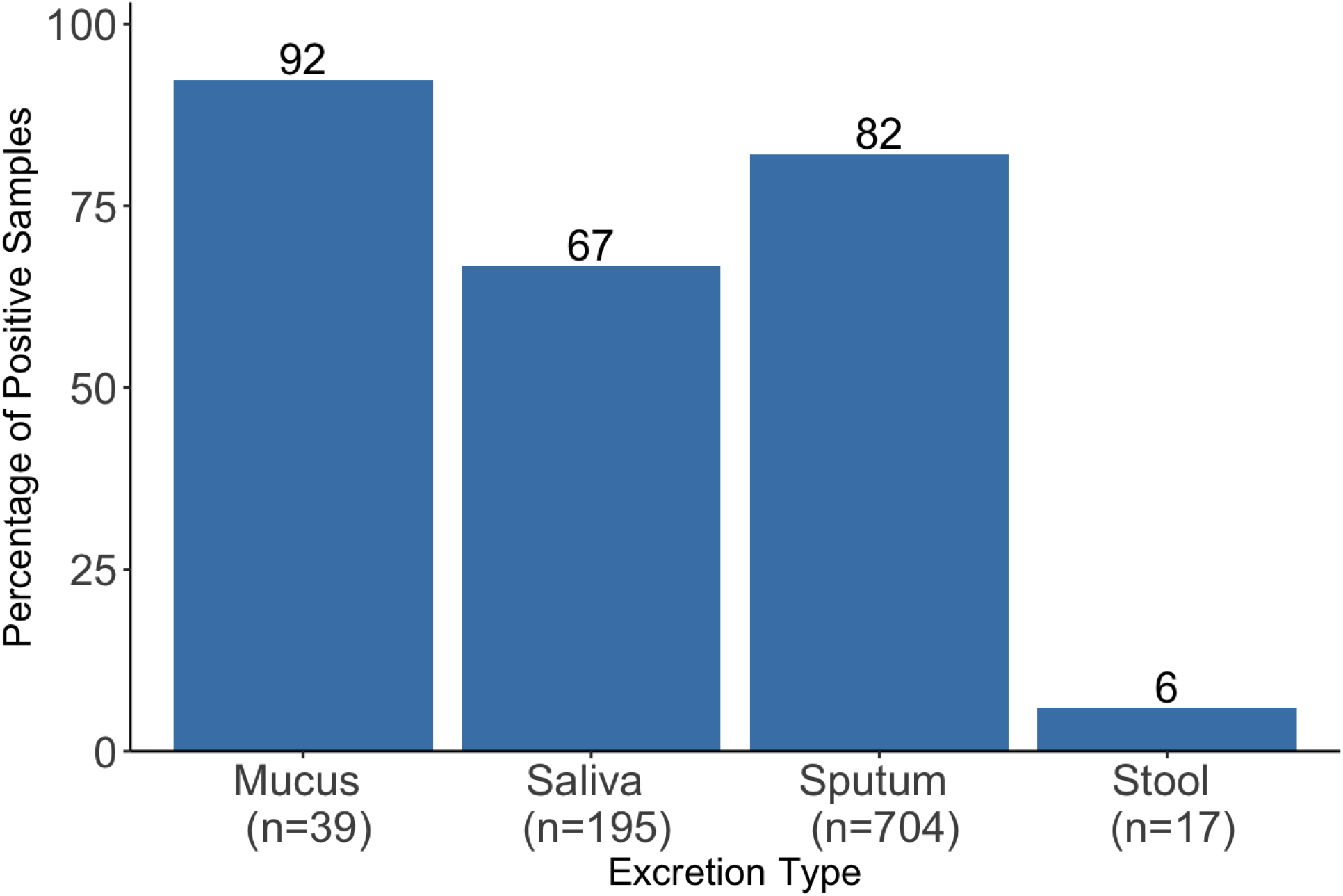
Weighted Average Percentage of Positive Rhinovirus Detections in Various Excretions of Subjects with Confirmed Rhinovirus Infections. The reported value for n is the total number of samples included in the meta-analysis for each excretion type. The number on top of each bar is the percent of samples positive. No data is reported for urine as no study measured rhinovirus in urine.

Since no urine data was identified in studies conducted with subjects with confirmed rhinovirus infections, we looked at data sets in which rhinovirus was tested in excretions of people with respiratory or gastrointestinal symptoms. None of these data sets reported on rhinovirus in urine (see Supporting Information Table S1).

#### Parainfluenza Virus

We identified 16 articles (15,19,20,22–34) that reported 32 data sets of parainfluenza virus 1, 2, 3, and/or 4 shedding in excretions in patients. These data sets all measured positivity rates in subjects with confirmed infections in cross-sectional studies that included between one (23–25,28) and 88 (26) subjects. No identified data sets reported parainfluenza virus concentrations in any excretion, and none reported longitudinal shedding data. None of the data sets measured parainfluenza virus in stool or urine. Thirty-one data sets measured parainfluenza virus using RT-PCR assays, and one data set used culture methods (19).

For the 32 data sets, some of them differentiated between the types of parainfluenza virus. Seven (7) data sets measured parainfluenza virus 1, six (6) measured parainfluenza virus 2, nine (9) measured parainfluenza virus 3, three (3) measured parainfluenza virus 4, and seven (7) did not specify a single type of parainfluenza virus. See Supporting Information Figure S1 for the weighted average positivity rate by parainfluenza type.

Positivity rates in mucus, saliva, and sputum are above 60% with the highest positivity rates observed in mucus (Figure 3). Fisher’s exact test resulted in a p-value of less than 0.001 rejecting the null hypothesis that positivity rates among excretions are the same.

**Figure 3.**
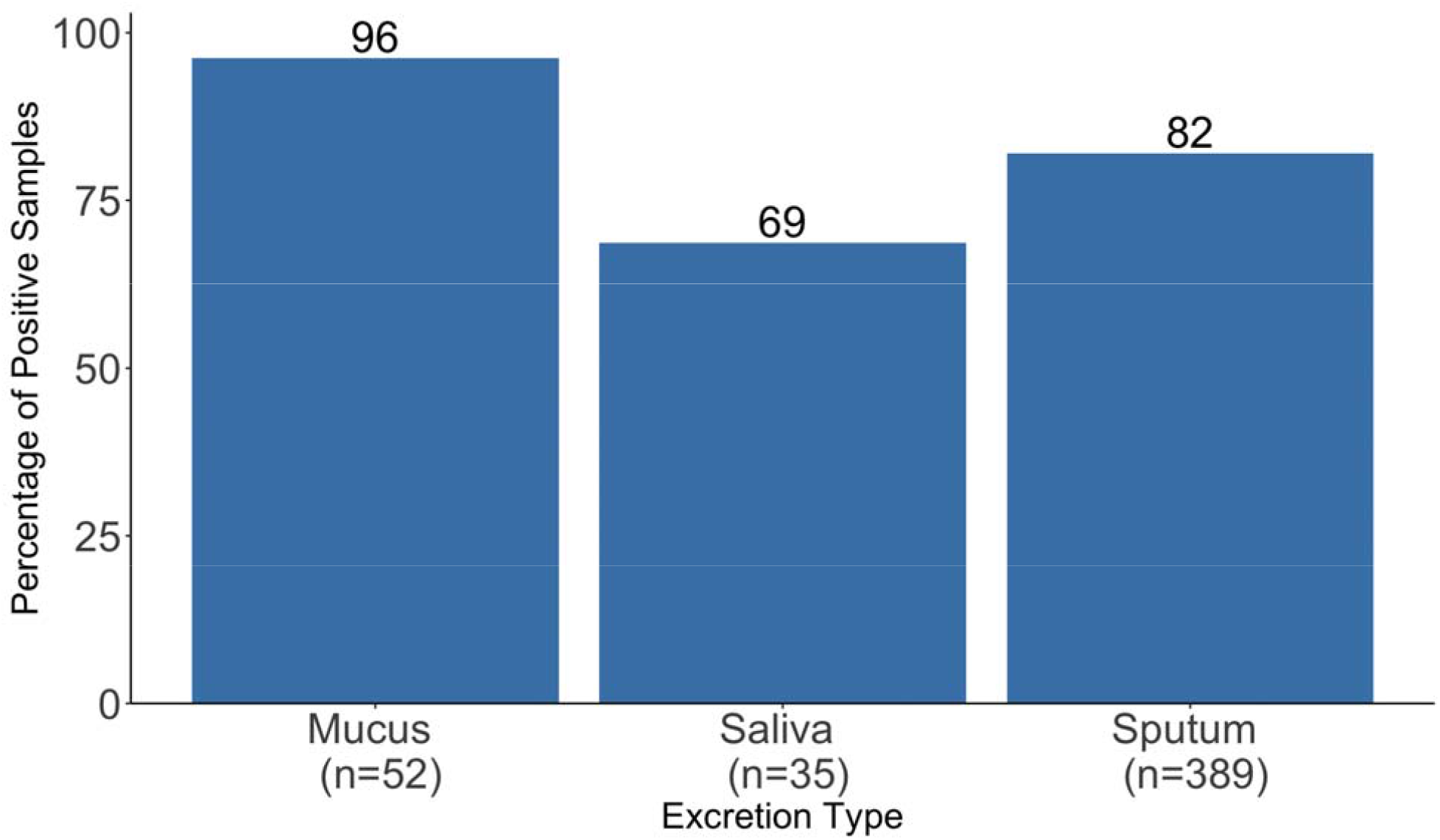
Weighted Average Percentage of Positive Parainfluenza Virus Detections in Various Excretions from Subjects with Confirmed Infection. The reported value for n is the number of tests included in the metaanalysis for each excretion type. No data is reported for stool or urine as no study measured parainfluenza virus in stool or urine. The number on top of each bar is the percent of samples positive.

**Figure 4:**
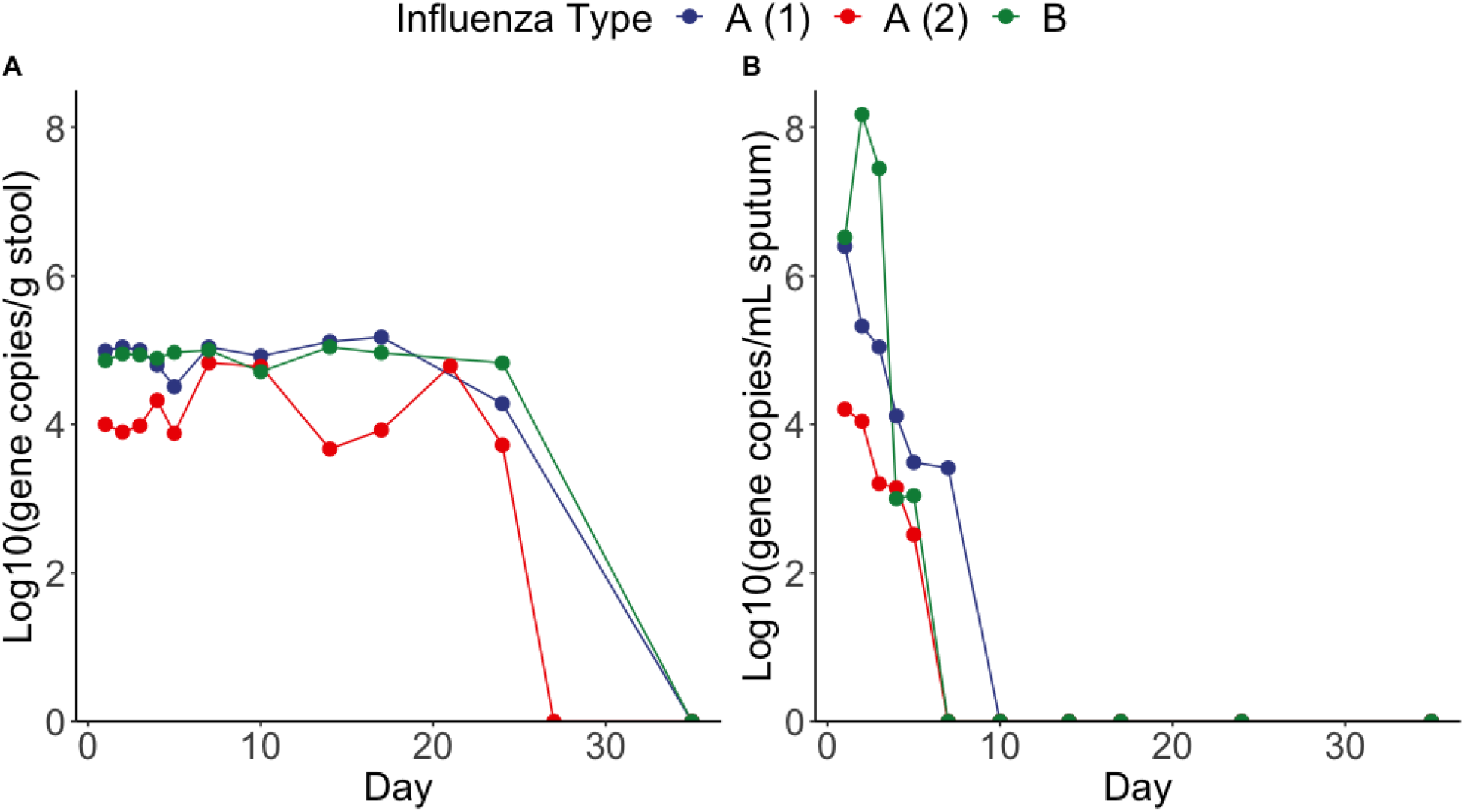
Concentrations of influenza measured in stool (A) and sputum (B) in three patients (two with Influenza A, as indicated by A (1) and A (2), and one with influenza B) over 35 days, as reported by Hirose et al. (44). A point located at a y-value of 0 represents ND (not detected), indicating the virus was not detected. Concentrations were reported in copies/mL for sputum and copies/g for stool.

Since no stool or urine data was identified in studies conducted with subjects with confirmed parainfluenza infections, we examined data sets that tested for parainfluenza virus in the excretions of people with respiratory or gastrointestinal symptoms. One of these data sets reported on parainfluenza virus in stool and reported 1/331 stool samples (0.3%) positive for parainfluenza virus 2, 3, or 4 (the study did not report data on parainfluenza virus 1) (35). None of the data sets measured parainfluenza virus in urine. See Supporting Information Table S2 for additional details of these data sets.

#### Influenza

We identified 37 articles (15,19,22–34,36–52,52–56) with data on influenza in patient excretions; these articles contained 77 influenza data sets. Seventy-six of the data sets were generated using RT-PCR and the remaining one used culture methods (19). Eight of the 77 data sets reported longitudinal shedding data (of these 8, two reported positivity rates (55), and six reported concentrations (44)), 10 reported concentration data from cross-sectional studies (38,40,41,44), and the remaining 59 reported cross-sectional positivity rate data from between one (24,28,45,46) and 120 (54) subjects with confirmed influenza infections.

The two longitudinal data sets reporting positivity rates (one for sputum, one for stool) came from a study that provided the number of days after influenza symptom onset that the excretion first tested positive for Influenza A H7N9: a median of 10.5 days (interquartile range 8.25 - 11.75) for sputum, and a median of seven days (interquartile range 7 - 7.75) for stool (55); overall 10/12 subjects were found to contain influenza A in at least one sputum sample and 6/12 subjects were found to contain influenza A in at least one stool sample.

The six longitudinal concentration data sets came from a single study (44) that examined the sputum and stool of three patients, two infected with influenza A H3N2 and one with influenza B, over 35 days (Table 3). In these patients, influenza was detected in stool through day 24 but was only detected in sputum through day five or 7. Sputum concentrations ranged from 3.3 × 10^2^ copies/mL to 1.5 × 10^8^ copies/mL, while stool concentrations ranged from 4.7 × 10^3^ copies/g to 1.0 × 10^5^ copies/g over time.

**Table 3.**
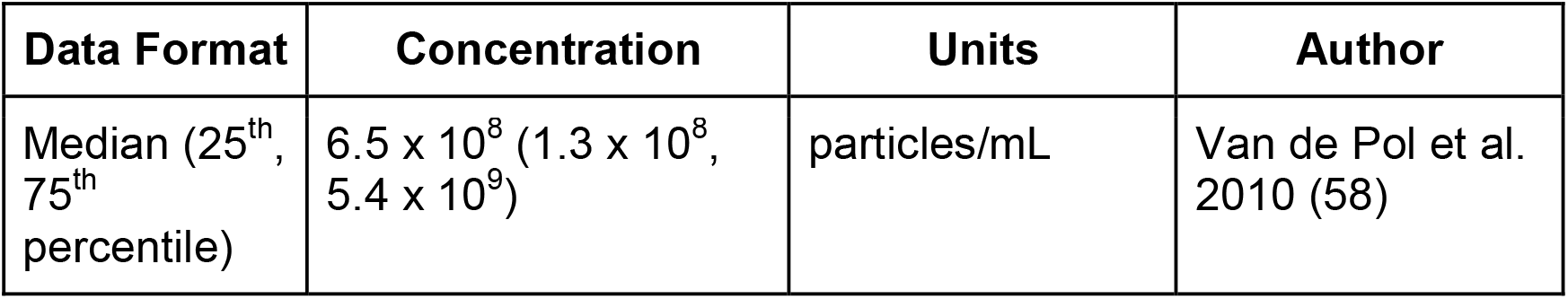
Measured concentrations of RSV in mucus.

| Data Format | Concentration | Units | Author |
| --- | --- | --- | --- |
| Median (25<sup>th</sup>, 75<sup>th</sup> percentile) | $6.5 \times 10^8$ ($1.3 \times 10^8$, $5.4 \times 10^9$) | particles/mL | Van de Pol et al. 2010 (58) |

Of the 69 cross-sectional data sets, 42 measured influenza A, 21 measured influenza B, and six did not discern between influenza A and B (19,25,29,30,33,47) (either because they did not specify which they measured, they measured both types with the same assay, or they measured both types with separate assays but reported combined results). The breakdown by type A or B is shown in the concentration data in Figure 5 and in the positivity rate data in Supplementary Information Figure S2. Some of the cross-sectional data sets additionally specified the influenza A subtype measured: seven measured influenza A H1N1 (39,41,48,50,52,53), 9 measured influenza A H3N2 (37,41,44), and two measured influenza A H7N9 (56).

**Figure 5.**
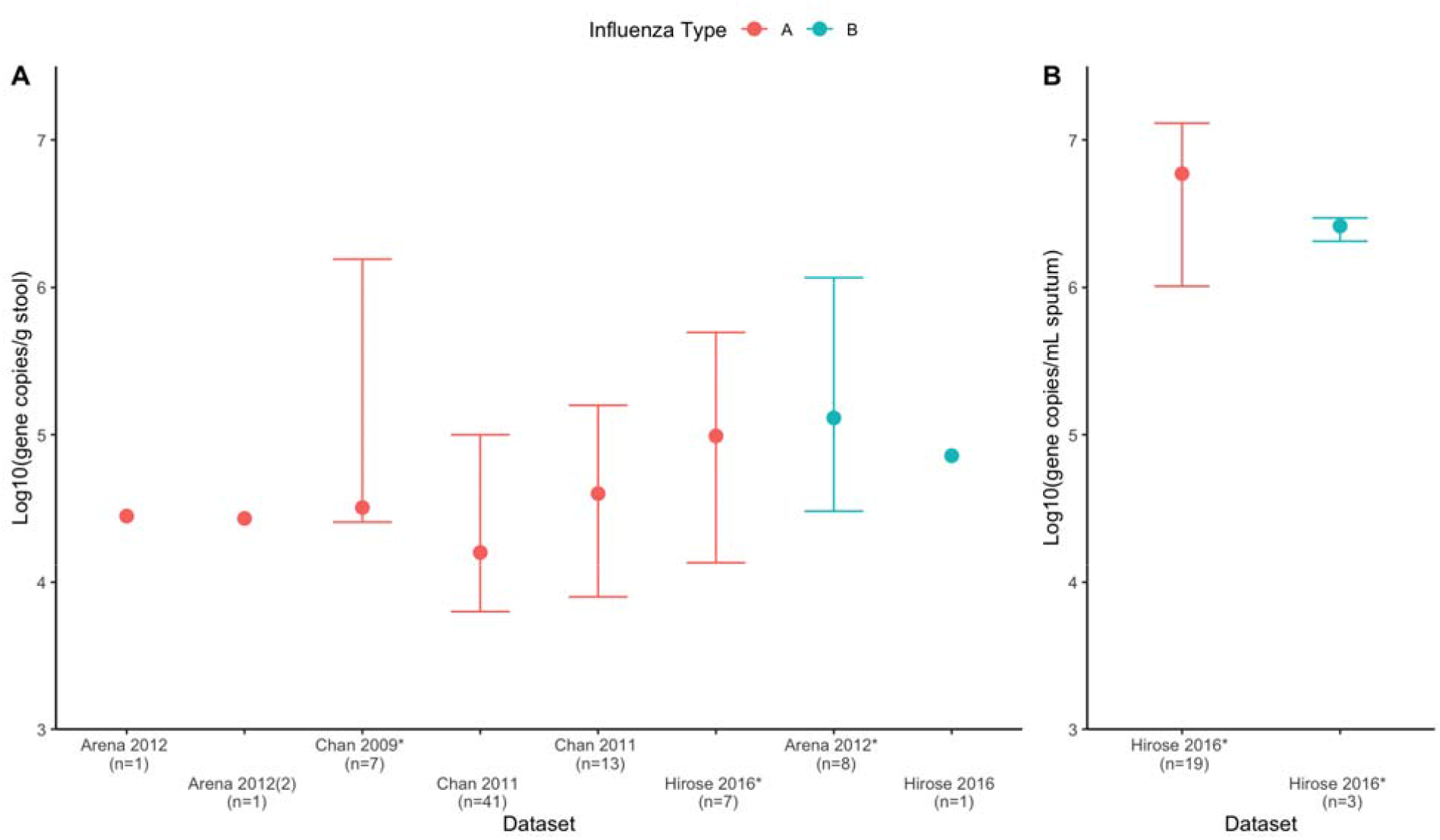
Concentrations of influenza in stool (A) and sputum (B), where points represent medians and bars indicate interquartile range (IQR) (single points indicate only one datapoint reported from data set by study authors). Data in (A) and (B) comes from only cross-sectional studies. Red points represent Influenza A and blue points represent Influenza B. The first author and year is used only to abbreviate the paper author list due to lack of space. There are two data sets from the same paper with the same number of subjects (n); these data sets are delineated by adding a (2) after one of the data sets. The number of subjects (n) for each data set is provided under the data set name. The papers from which these data sets came include Arena et al. (38), Chan et al. (40), Chan et al. (41), and Hirose et al. (44). The * next to the first author and year indicates datasets that display summary statistics that we calculated.

Of the 10 data sets reporting influenza concentrations measured in cross-sectional studies, eight measured the concentration of influenza in stool (reported and calculated medians span 1.6 × 10^4^ to 1.3 × 10^5^ copies/g stool), and two measured the concentration of influenza in sputum (reported and calculated medians span 2.6 × 10^6^ to 5.9 × 10^6^ copies/mL sputum) (Figure 5). Samples were acquired from patients at a variety of times post symptom onset (38,40,41,44). Concentrations were measured using RT-PCR methods. Authors provided summary statistics and point values for 5 datasets (38,41,44), and summary statistics were calculated using data extracted via WebPlotDigitizer (12) for the remaining 5 datasets (38,40,44).

Figure 6 shows the weighted average positivity rate data from cross-sectional studies separated by excretion type. Positivity rates of mucus, saliva, and sputum were greater than 80% while positivity rates for urine and stool were 58% and 36%, respectively. A chi-square test resulted in a p-value of less than 0.001, rejecting the null hypothesis that positivity rate is the same across excretions. See Supporting Information Table S3 for data sets of subjects without confirmed influenza infections.

**Figure 6.**
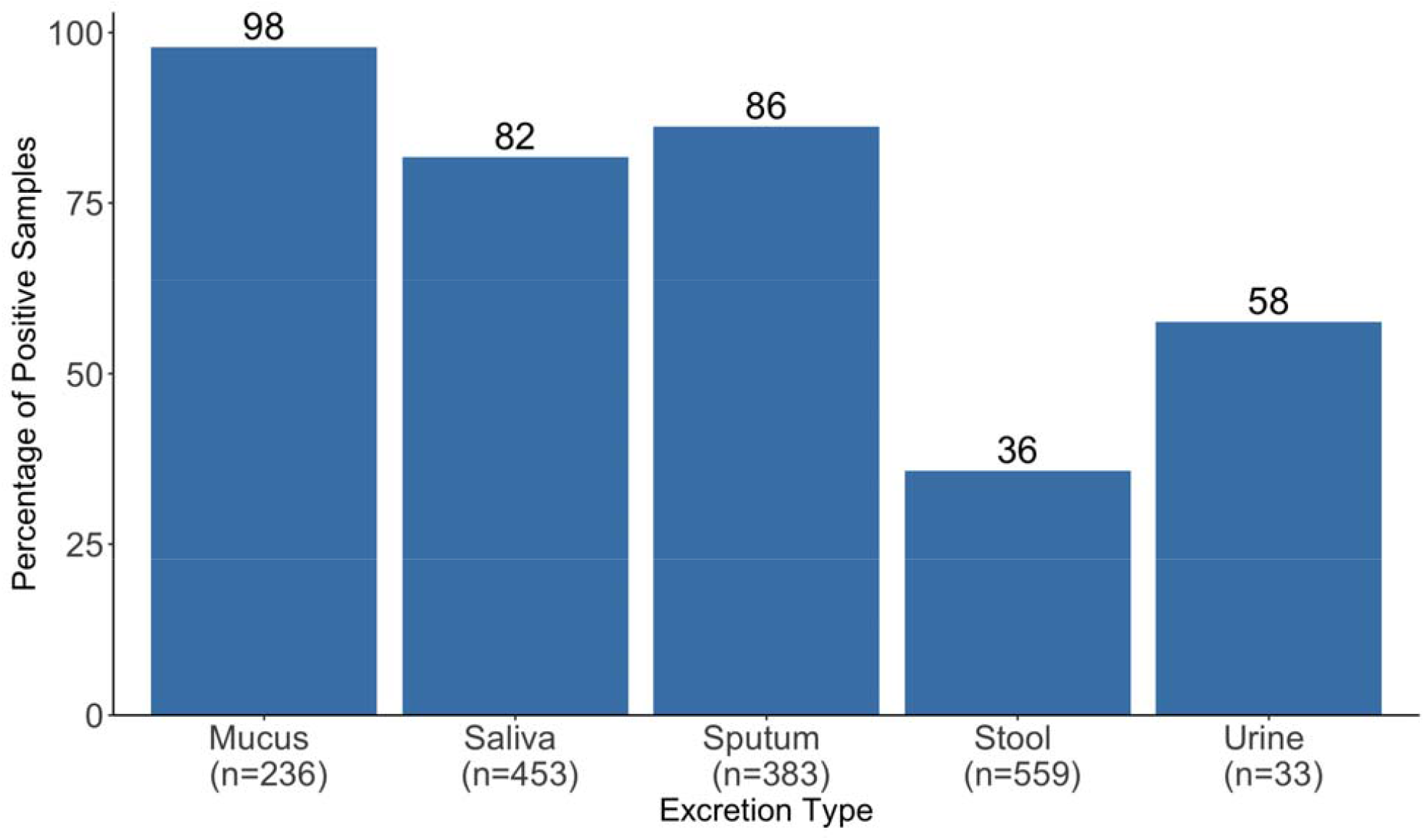
Weighted Averages for Percentage of Positive Influenza Virus A or B Detections in Various Excretions from Subjects with Confirmed Infection. The reported value for n is the number of samples included in the weighted average for each excretion type. The number on top of each bar is the percent of samples positive.

#### Respiratory Syncytial Virus (RSV)

We identified 20 studies containing 28 data sets with information on the shedding of RSV in excretions of subjects. One data set reported longitudinal positivity rates, one data set reported concentrations of RSV in excretions from a cross-sectional study (specifically in mucus, Table 3), and the remaining 26 reported positivity rate data from cross-sectional studies with between one (23) and 323 (26) subjects with confirmed RSV infections. Two data sets measured RSV A, three data sets measured RSV B, and the remaining 23 did not specify a type of RSV in their analyses. Twenty-eight of the data sets were generated using RT-PCR and the remaining one used culture methods (19).

The longitudinal data set reported the percent of RSV detections in mucus from 38 subjects over 27 days, with RSV detected in 38/38 (100%) of subjects on day 0, 20/38 (53%) of subjects on days 5-13, 9/34 (26%) of subjects on days 12-20, and 7/34 (21%) on days 18-27 (57).

One data set (58) measured RSV concentrations in 138 samples of mucus from 31 mechanically ventilated infants (Table 3) in a cross sectional study. Concentrations were measured using RT-PCR. The authors of the study use the term “particles” in reporting units, which is assumed to mean virus. Summary statistics were calculated from data extracted using WebPlotDigitizer (12).

Figure 7 shows the weighted average positivity rate for each excretion in subjects with confirmed RSV infections, as calculated from the 26 cross-sectional positivity rate data sets. A breakdown of RSV A versus RSV B positivity rates is shown in Supporting Information Figure S3. While the weighted averages for mucus, saliva, and sputum are all >75%, the weighted average for stool is 14% and there were no urine samples positive for RSV. A chi-square test resulted in a p-value of less than 0.001, rejecting the null hypothesis that positivity rates are the same across excretions.

**Figure 7.**
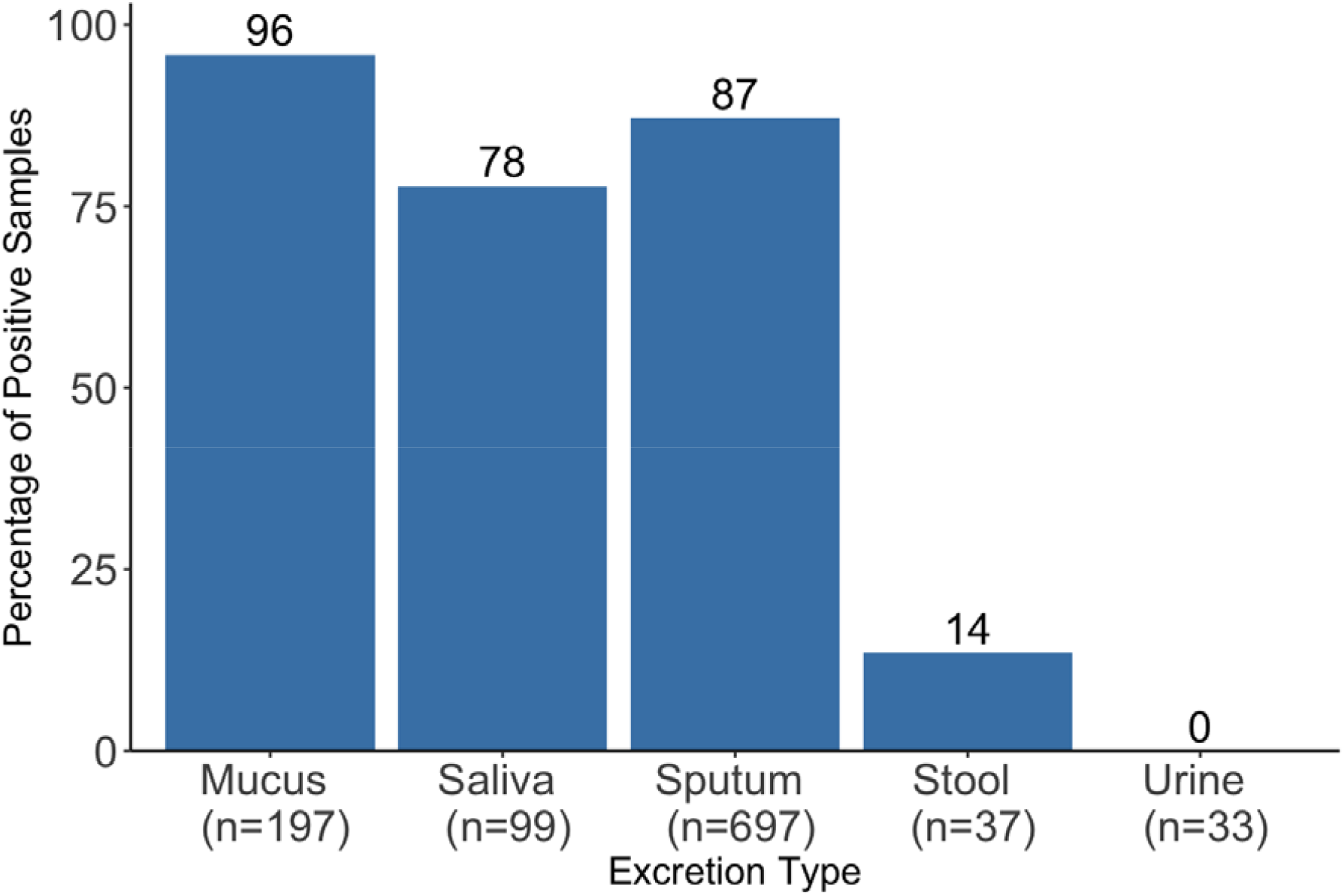
Weighted Average Percentage of Positive RSV Detections in Various Excretions from Subjects with Confirmed Infection. The reported value for n is the number of samples included in the metaanalysis for each excretion type. The number on top of each bar is the percent of samples positive.

Because RSV was not detected in urine in any subject, we examined data sets that assayed for RSV in the urine of subjects with respiratory or gastrointestinal symptoms. No identified study tested for the presence of RSV in urine. See Supporting Information Table S4 for a list of these data sets.

#### Metapneumovirus

We identified 15 studies (15,20,24–32,34,46,57,59) containing 20 data sets with information on the shedding of metapneumovirus in excretions of subjects with confirmed metapneumovirus infections. One data set reported longitudinal positivity rates, while the remaining 19 reported positivity rate data from cross-sectional studies with between one (20,29,46) and 156 (26) subjects. All 20 data sets measured the virus using RT-PCR-based methods.

The longitudinal data set reported the positivity rates of metapneumovirus detection in mucus of seven subjects over 24 days, with 7/7 (100%) testing positive on day 0 since diagnosis with metapneumovirus via nasopharyngeal aspirate sample, 3/7 (43%) positive on days 4-10, 0/7 positive on days 12-17, and 0/6 positive on days 20-24 (57).

Figure 8 shows the weighted average metapneumovirus positivity rates in various excretions from the 19 cross-sectional data sets of subjects with confirmed metapneumovirus infection. Mucus and sputum have the highest weighted averages at >80%, and stool and urine have the lowest (0%), each being evaluated by only a single data set with data from six subjects that did not detect any metapneumovirus. A Fisher’s exact test resulted in a p-value of less than 0.001, rejecting the null hypothesis that positivity rates are the same across excretions.

**Figure 8.**
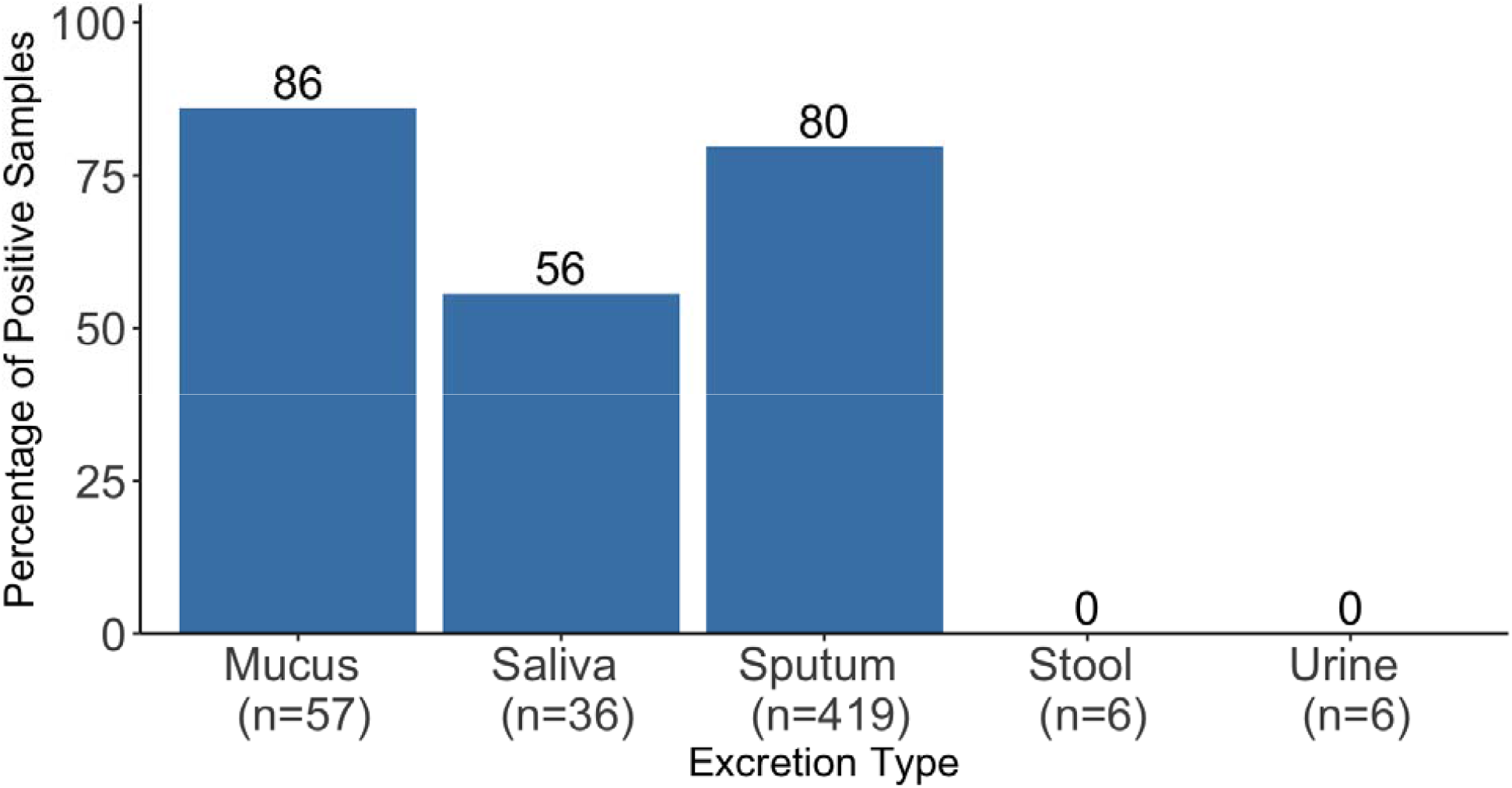
Weighted Average Percentage of Positive Metapneumovirus Detections in Various Excretions from Subjects with Confirmed Metapneumovirus Infection. The reported value for n is the number of samples included in the metaanalysis for each excretion type. The number on top of each bar is the percent of samples positive.

Since metapneumovirus was not detected in any stool or urine samples of subjects with confirmed metapneumovirus infections, we examined data sets testing for metapneumovirus in the excretions of subjects with respiratory or gastrointestinal symptoms. Metapneumovirus was detected in the stool of subjects with respiratory symptoms, where it was found in 2/331 (0.6%) stool samples (35). However, no identified data sets tested urine for metapneumovirus. See Supporting Information Table S5 for additional information on these data sets.

#### Seasonal Coronaviruses

We identified 16 studies (15,17,20,22,23,25–28,30,31,34,46,60–62) containing 43 data sets on the shedding of human coronaviruses 229E, HKU1, NL63, and/or OC43 in various excretions. No data sets reported any longitudinal shedding data, and no study reported concentration data. All identified data sets took the form of positivity rates, reporting cross-sectional data with sample sizes ranging from one (20,22,28,34,60) to 62 (62) subjects with confirmed seasonal coronavirus infection. Of the 43 data sets, 42 measured the virus using RT-PCR-based methods while one used both cultivation and RT-PCR-based methods (61).

Figure 9 shows the weighted average positivity rates of seasonal coronaviruses in various excretions from the 43 cross-sectional data sets of subjects with confirmed seasonal coronavirus infection. Sputum and saliva both have the highest positivity rates (86% and 75%, respectively) and stool and mucus have the lowest (39% and 25%, respectively). No data sets of positivity rates of seasonal coronaviruses in urine of subjects with confirmed seasonal coronavirus infection were identified in this review. A Fisher’s exact test resulted in a p-value of less than 0.001, rejecting the null hypothesis that positivity rates are the same across excretions. Supporting Information Figure S4 shows the weighted averages separated by seasonal coronavirus type.

**Figure 9.**
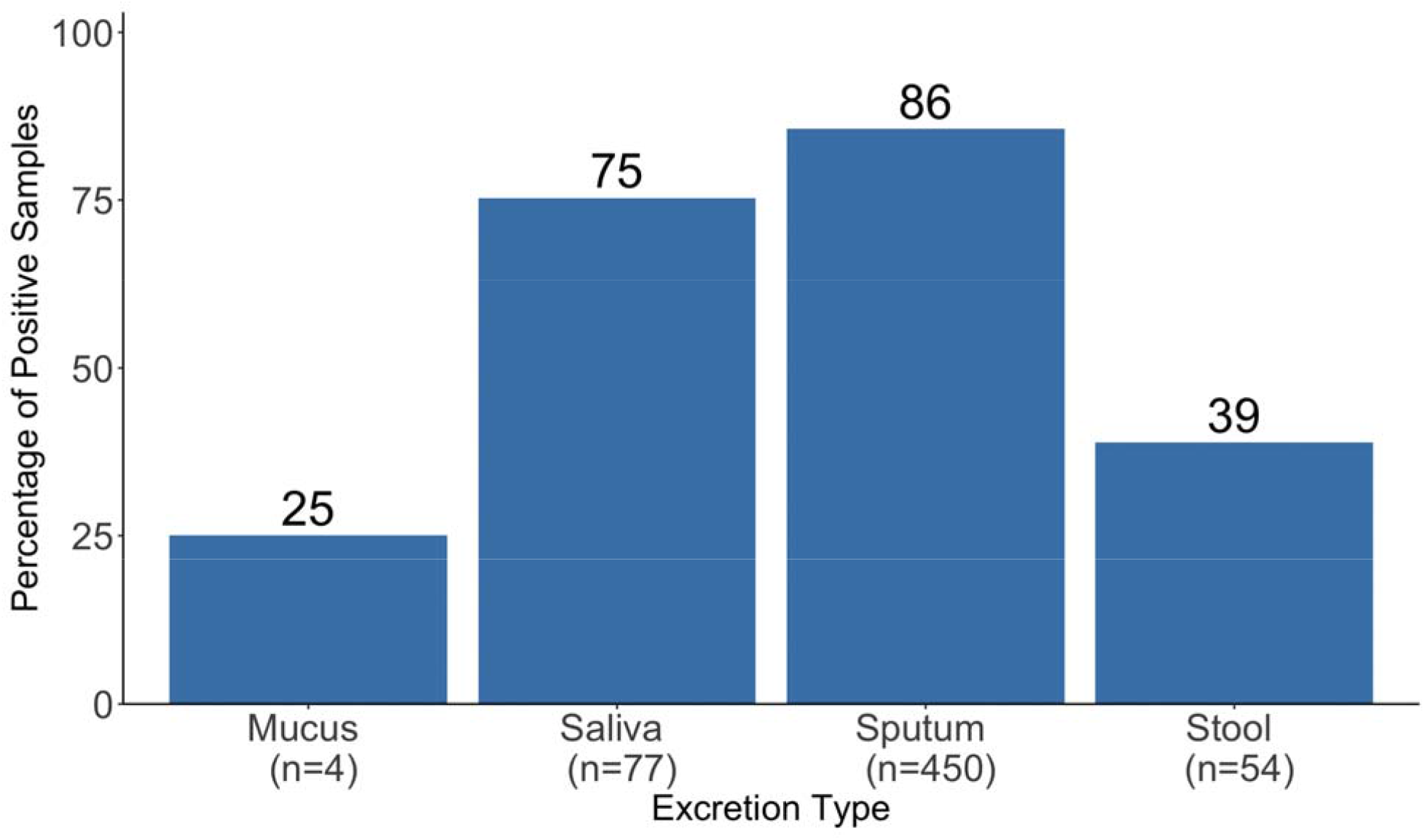
Weighted Averages of Percentage of Positive Seasonal Coronavirus Detections in Various Excretions from Subjects with Confirmed Seasonal Coronavirus Infection. The reported value for n is the number of samples included in the metaanalysis for each excretion type. The number on top of each bar is the percent of samples positive.

Since human coronaviruses were not measured in urine of subjects with confirmed infections, we examined data sets that tested for seasonal coronaviruses in the excretions of subjects with respiratory or gastrointestinal symptoms. No identified data sets measured seasonal coronaviruses in urine. See Supporting Information Table S6 for additional information on these data sets.

## Discussion

There are limited data available on the shedding of respiratory viruses in stool, saliva, sputum, mucus, and urine. Our systematic review of the literature found that the vast majority of data sets characterize positivity rates of respiratory viruses in excretions (195/220), while only a few quantify respiratory virus concentration in excretions in a cross-sectional study design (14/220) and even fewer examine longitudinal shedding patterns in excretions (11/220). Additionally, the majority of data sets evaluated respiratory viruses in saliva, sputum, and mucus. There was less data available for respiratory virus detection in stool, as no data sets were available for parainfluenza virus and the one data set that tested for metapneumovirus in stool failed to detect any (although one study examining subjects with respiratory or gastrointestinal symptoms reported positive detections of both parainfluenza virus and metapneumovirus in stool (35)). Data sets on respiratory viruses in urine are even more sparse; with no studies examining rhinovirus, parainfluenza virus, and seasonal coronaviruses in urine and limited work examining RSV and metapneumovirus in urine (1 study assayed both RSV and metapneumovirus in urine but failed to detect either virus (57)). Only influenza has been detected in the urine of subjects with the respective diagnosed infection. This emphasizes the finding that respiratory viruses have been characterized in different excretions to differing extents, limiting the current knowledge of viral concentrations in stool and urine, the more understudied excretions.

Data availability also differed according to respiratory virus type. There were the most excretion data sets for influenza (n = 75) and the least for metapneumovirus and rhinovirus (n = 20 for each). Table 4 highlights the uneven distribution of data as shown in the number of samples evaluated in each excretion and virus.

**Table 4.** Summary of positivity rates, reported as percentage (number of samples positive/number of total samples), of respiratory viruses in excretions of subjects with confirmed infections. 0 indicates excretions were tested but none were positive for the presence of the virus, while n/a indicates no excretions were tested for the presence of the virus. * indicates the virus was not detected in subjects with confirmed infection but has been detected in the excretion although in patients with symptoms. RV is rhinovirus, PIV is parainfluenza virus, Flu is influenza, RSV is respiratory syncytial virus, HMPV is human metapneumovirus, and HCoV is human seasonal coronavirus.

|  | RV | PIV | Flu | RSV | HMPV | HCoV |
| --- | --- | --- | --- | --- | --- | --- |
| Mucus | 92% | 96% | 98% | 96% | 86% | 25% |
|  | (36/39) | (50/52) | (231/236) | (189/197) | (49/57) | (1/4) |
| Saliva | 67%<br>(130/195) | 69%<br>(24/35) | 82%<br>(370/453) | 78%<br>(77/99) | 56%<br>(20/36) | 75%<br>(58/77) |
| Sputum | 82%<br>(578/704) | 82%<br>(319/389) | 86%<br>(330/383) | 87%<br>(608/697) | 80%<br>(334/419) | 86%<br>(385/450) |
| Stool | 6%<br>(1/17) | n/a* | 36%<br>(200/559) | 14%<br>(5/37) | 0%*<br>(0/6) | 39%<br>(21/54) |
| Urine | n/a | n/a | 58%<br>(19/33) | 0%<br>(0/33) | 0%<br>(0/6) | n/a |

Across the five different excretions, positivity rates for each virus were highest in mucus, followed by sputum and then saliva (summarized in Table 4), and hypothesis testing rejected the null hypothesis that positivity rates were the same across excretions. The positivity rate of human coronavirus in mucus is the only exception to this pattern, but data were limited (only four samples of mucus tested for human coronaviruses compared to between 39 and 236 mucus samples tested for the other viruses).

There were very few quantitative measurements of respiratory virus concentrations in excretions. The most concentration data were available for influenza for which there were 21 data sets (6 longitudinal and 15 cross-sectional), while no concentrations of parainfluenza virus, metapneumovirus, or seasonal coronaviruses were reported in any excretions. No data sets measured any respiratory virus concentration in urine. Comparing viral concentrations between excretions and between respiratory viruses is challenging because of the diversity of quantification methods used and units reported. Units reported include TCID_50_/g (culture-based methodology), and genomes/mg, copies/g, and particles/mL (RT-PCR-based methodology). Some studies that reported quantitative-type data on respiratory viruses in excretions were excluded from this review because they quantified viruses using units that were not externally valid (e.g. CT values from real time PCR machines) or failed to report sufficient methods to determine the concentration of the respiratory virus in the excretion (e.g. concentration in copies/mL of viral transport medium).

This work reveals that there is a significant knowledge gap on respiratory virus concentrations in excretions. This information is needed to link concentrations of respiratory viruses in wastewater to the number of people infected with the virus in the population contributing to wastewater. We recommend that future studies should explicitly characterize concentrations of respiratory viruses in mucus, saliva, sputum, stool, and urine, reporting standard units (e.g. copies per unit volume of excretion) and complete methods including details established by the Minimum Information for Publication of Quantitative Real-Time PCR Experiments (MIQE) (63) and Environmental Microbiology Minimum Information (EMMI) Guidelines (64). Data are needed across all respiratory virus and excretion types to build more robust data sets, but are especially needed for parainfluenza virus, metapneumovirus, and human coronavirus where there was no concentration data identified in this review. While positivity rate data sets are useful in estimating the proportion of infected people that shed a certain respiratory virus in excretions, they cannot provide quantitative estimates of viral shedding necessary to begin to translate wastewater concentrations to individual case numbers in a community. Additionally, longitudinal studies that examine the concentrations of respiratory viruses in excretions over time are critical in providing information on both the duration of shedding and how concentrations of viruses in excretions vary over time. Such data, for example, are needed to use wastewater data to predict effective reproductive numbers of pathogens (65). Very few studies identified in this review reported longitudinal data and only one study on influenza reported longitudinal concentration data. Additional factors to consider in predicting effective reproductive numbers of pathogens through wastewater include the relative contribution of each excretion to wastewater, the design of the sewer system, the wastewater sampling location, and the sample matrix, among others (65). Public health officials and modelers interested in applying these results to quantitatively link their wastewater concentrations to numbers of infected individuals should be aware of these limitations and data gaps.

There are several limitations to this review. We only included studies written in English, which could have excluded studies with relevant data. We did not perform any formal evaluations of bias across the included studies. Additionally, we chose to focus on data sets with measurements made in direct excretions, excluding samples from areas of the body that generate excretions (e.g. swabs and lavages). This narrowed the body of literature applicable to this review. We also focused on positivity rates in the excretions of subjects with confirmed infections, rather than subjects whose infection status was unknown, again narrowing the applicable body of literature. This decision was made because positivity rates from subjects whose infection status is unknown are not useful in determining the shedding rates of viruses in excretions; rather the only information relevant to this review that they can provide is evidence that respiratory viruses can be shed in certain excretions. Some cross-sectional data sets inadvertently sampled from the same subject multiple times, which could impact the positivity rate data. The chosen statistical tests we performed (chi-square test and Fisher’s exact test) do not account for correlation, but some of our data was correlated. However, we could not account for this correlation when comparing the data across all excretion types. Finally, we were unable to perform a formal meta-analysis on concentration data given the limited data and inconsistent units reported in the concentration data. As such, we could not examine whether shedding patterns were different across different viral subtypes (for example, RSV A versus RSV B).

## Supporting information

Prisma Checklist

## Data Availability

All data used in this study is available publicly through the Stanford Data Repository.

https://doi.org/10.25740/vj779wy5347

## Author Contributions

S.L. Conceptualization, Data Curation, Formal analysis, Investigation, Methodology, Visualizations, Writing – Original Draft Preparation, Writing – Review & Editing. A.B.B. Conceptualization, Funding Acquisition, Methodology, Project Administration, Resources, Supervision, Validation, Writing – Original Draft Preparation, Writing – Review & Editing. M.K.W. Conceptualization, Writing - Review & Editing, Methodology.

