## Supporting Information for "Respiratory virus concentrations in human excretions that contribute to wastewater: A systematic review"

Table S1. Reported Positivity Rates of Rhinovirus in Stool for Subjects with Respiratory or Gastrointestinal Symptoms (1–7)

| **Symptom Type** | **Positivity Rate (Percentage)** | **Author** |
| --- | --- | --- |
| Gastrointestinal | 11/164 (6.7%) | Bergallo et al. 2019 |
| Gastrointestinal | 39/372 (10.5%) | Harvala et al. 2012 |
| Gastrointestinal | 30/1294 (2.3%) | Khoonta et al. 2017 |
| Gastrointestinal | 58/734 (7.9%) | Lau et al. 2012 |
| Respiratory | 13/331 (3.9%) | Minodier et al. 2017 |
| Gastrointestinal | 36/689 (5.2%) | Rovida et al. 2013 |
| Respiratory and/or gastrointestinal | 149/425 (35.1%) | Savolainen-Kopra et al. 2013 |

Table S2. Reported Positivity Rates of Parainfluenza for Subjects with Respiratory Symptoms (5,8)

| **Excretion Type** | **Positivity Rate (Percentage)** | **Author** |
| --- | --- | --- |
| Mucus | 3/46 (6.5%) | Huang et al. 2020 |
| Sputum | 48/1285 (3.7%) | Huang et al. 2020 |
| Stool | 1/331 (0.3%) | Minodier et al. 2017 |

Table S3. Reported Positivity Rates of Influenza in Subjects with Gastrointestinal or Respiratory Virus Symptoms (5,8–12)

| **Author** | **Influenza Type** | **Symptom Type** | **Excretion Type** | **Positivity Rate (Percentage)** |
| --- | --- | --- | --- | --- |
| Arena et al. 2012 | A H3N2 | Gastrointestinal | Stool | 1/138 (0.7%) |
| Arena et al. 2012 | A H1N1 | Gastrointestinal | Stool | 1/138 (0.7%) |
| Arena et al. 2012 | B | Gastrointestinal | Stool | 8/138 (5.8%) |
| Huang et al. 2020 | A or B | Respiratory | Mucus | 6/46 (13%) |
| Huang et al. 2020 | A or B | Respiratory | Sputum | 129/1285 (10%) |
| Minodier et al. 2017 | A | Respiratory | Stool | 12/331 (3.6%) |
| Minodier et al. 2017 | B | Respiratory | Stool | 13/331 (3.9%) |
| Covalciuc et al. 1999 | A or B | Respiratory | Mucus | 49/79 (62%) |
| Covalciuc et al. 1999 | A or B | Respiratory | Sputum | 46/70 (65.7%) |
| Ye et al. 2018 | A | Respiratory | Sputum | 2301/26466 (8.7%) |
| Ye et al. 2018 | B | Respiratory | Sputum | 2912/26466 (11%) |
| Xie et al. 2020 | A or B | Gastrointestinal | Stool | 13/440 (3%) |

Table S4. Reported Positivity Rates of RSV for Subjects with Respiratory Symptoms (5,8,11,13)

| **Excretion Type** | **Positivity Rate (Percentage)** | **Author** |
| --- | --- | --- |
| Sputum | 23/589 (4%) | Cattoir et al. 2019 |
| Mucus | 2222/11815 (19%) | Cattoir et al. 2019 |
| Sputum | 37/1285 (3%) | Huang et al. 2020 |
| Mucus | 0/46 (0%) | Huang et al. 2020 |
| Stool | 0/331 (0%) | Minodier et al. 2017 |
| Sputum | 3326/26466 (13%) | Ye et al. 2018 |

Table S5. Reported Positivity Rates of Metapneumovirus for Subjects with Respiratory Symptoms (5,8,13)

| **Excretion Type** | **Positivity Rate (Percentage)** | **Author** |
| --- | --- | --- |
| Sputum | 21/589 (4%) | Cattoir et al. 2019 |
| Mucus | 960/11815 (8%) | Cattoir et al. 2019 |
| Sputum | 22/1285 (2%) | Huang et al. 2020 |
| Mucus | 0/46 (0%) | Huang et al. 2020 |
| Stool | 2/331 (1%) | Minodier et al. 2017 |
| Mucus | 9/331 (3%) | Minodier et al. 2017 |

Table S6. Reported Positivity Rates of Seasonal Coronaviruses in Subjects with Gastrointestinal or Respiratory Virus Symptoms (5,6,8,14–17)

| **Symptom Type** | **Coronavirus Type** | **Excretion Type** | **Positivity Rate (Percentage)** | **Author** |
| --- | --- | --- | --- | --- |
| Gastrointestinal | 229E | Stool | 0/479 (0%) | Esper et al. 2010 |
| Gastrointestinal | OC43 | Stool | 0/479 (0%) | Esper et al. 2010 |
| Gastrointestinal | NL63 | Stool | 0/479 (0%) | Esper et al. 2010 |
| Gastrointestinal | HKU1 | Stool | 4/479 (1%) | Esper et al. 2010 |
| Respiratory | any | Sputum | 40/1285 (3%) | Huang et al. 2020 |
| Respiratory | any | Mucus | 2/46 (4%) | Huang et al. 2020 |
| Gastrointestinal | any | Stool | 6/218 (3%) | Jevsnik et al. 2016 |
| Respiratory | any | Stool | 4/331 (1%) | Minodier et al. 2017 |
| Gastrointestinal | OC43 | Stool | 10/878 (1%) | Risku et al. 2010 |
| Gastrointestinal | HKU1 | Stool | 6/878 (0.7%) | Risku et al. 2010 |
| Gastrointestinal | 229E | Stool | 2/878 (0.2%) | Risku et al. 2010 |
| Gastrointestinal | NL63 | Stool | 4/878 (0.5%) | Risku et al. 2010 |
| Gastrointestinal | any | Stool | 5/689 (0.7%) | Rovida et al. 2013 |
| Respiratory | 229E | Sputum | 2/202 (1%) | Vabret et al. 2001 |
| Respiratory | OC43 | Sputum | 4/202 (2%) | Vabret et al. 2001 |
| Respiratory | 229E | Mucus | 3/146 (2%) | Vabret et al. 2001 |
| Respiratory | OC43 | Mucus | 3/146 (2%) | Vabret et al. 2001 |

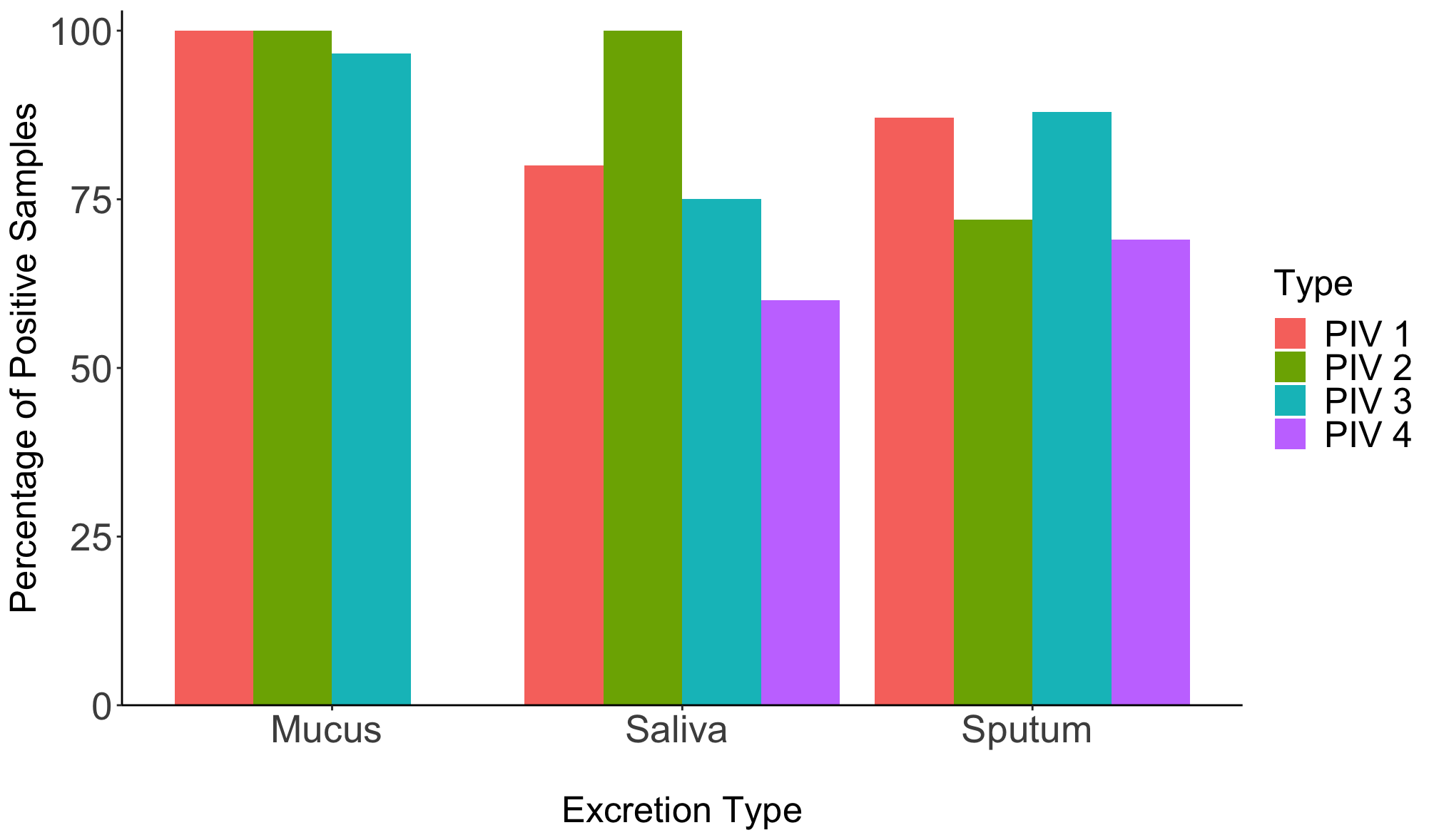

Figure S1. Weighted Average Percentage of Positive Parainfluenza Virus Detections in Various Excretions from Subjects with Confirmed Infection by Parainfluenza Virus (PIV) Type. Data sets that specified the measurement of one parainfluenza virus type were included (18–26); data sets that did not specify the measurement of one parainfluenza virus type were excluded. Breakdowns for n are as follows: Mucus: PIV1(n=8), PIV2(n=6), PIV3(n=28), PIV4(n=0), Saliva: PIV1(n=5), PIV2(n=10), PIV3(n=4), PIV4(n=5), Sputum: PIV1 (n=101), PIV2(n=25), PIV3(n=91), PIV4(n=42). Graphics were created using the statistical computing program R (27).

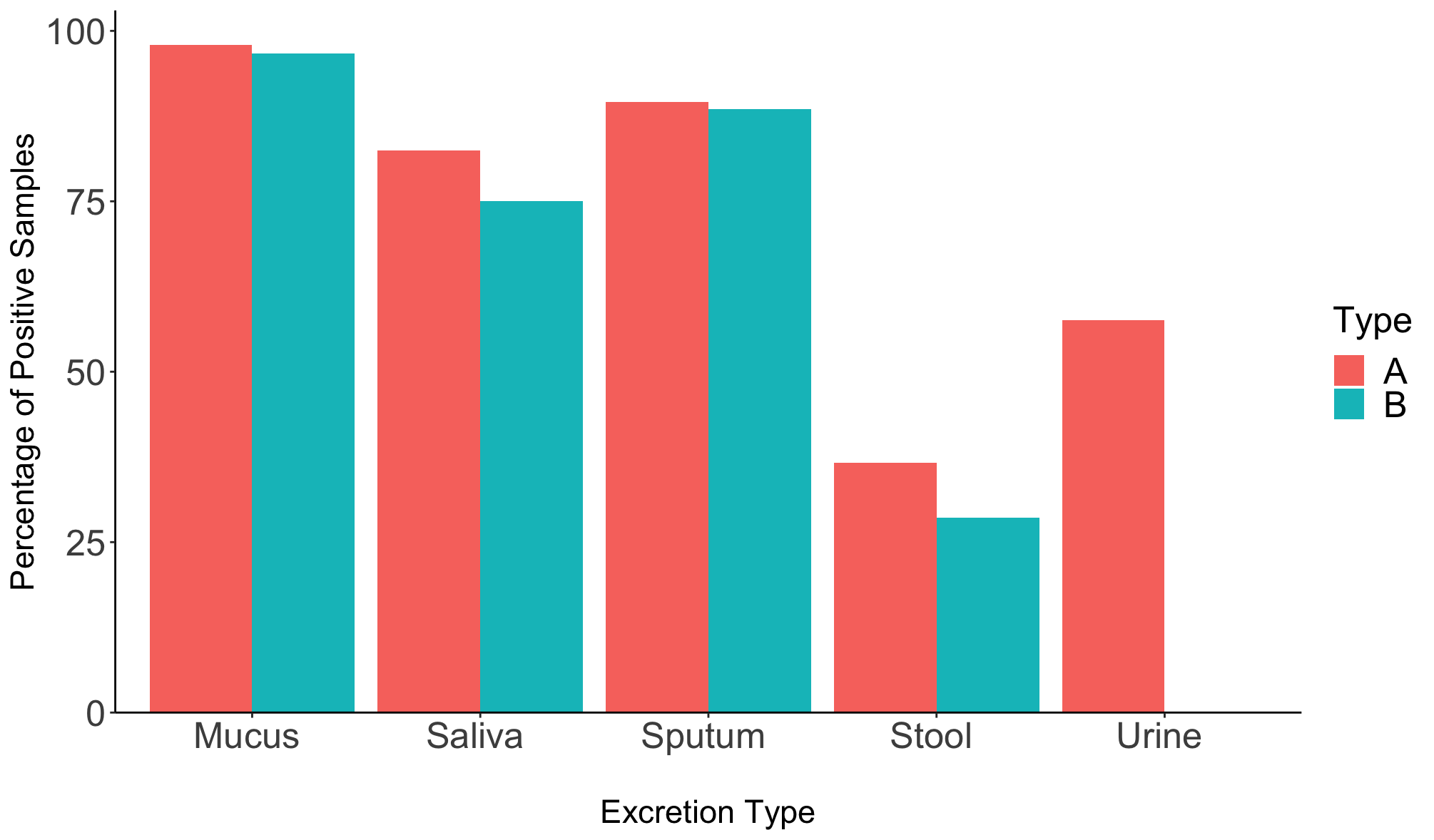

Figure S2. Weighted Averages of Percentage of Positive Influenza Virus A or B Detections Separated by Influenza Type in Various Excretions from Subjects with Confirmed Infection. Data sets that specified the measurement of one parainfluenza virus type were included (19–22,24–26,28–49); data sets that did not specify the measurement of one influenza type were excluded. Breakdowns for n are as follows: Mucus: A(n=194), B(n=30), Saliva: A(n=361), B(n=56) Sputum: A(n=279), B(n=61) Stool: A(n=494), B(n=63), Urine: A(n=33), B(n=0). Graphics were created using the statistical computing program R (27).

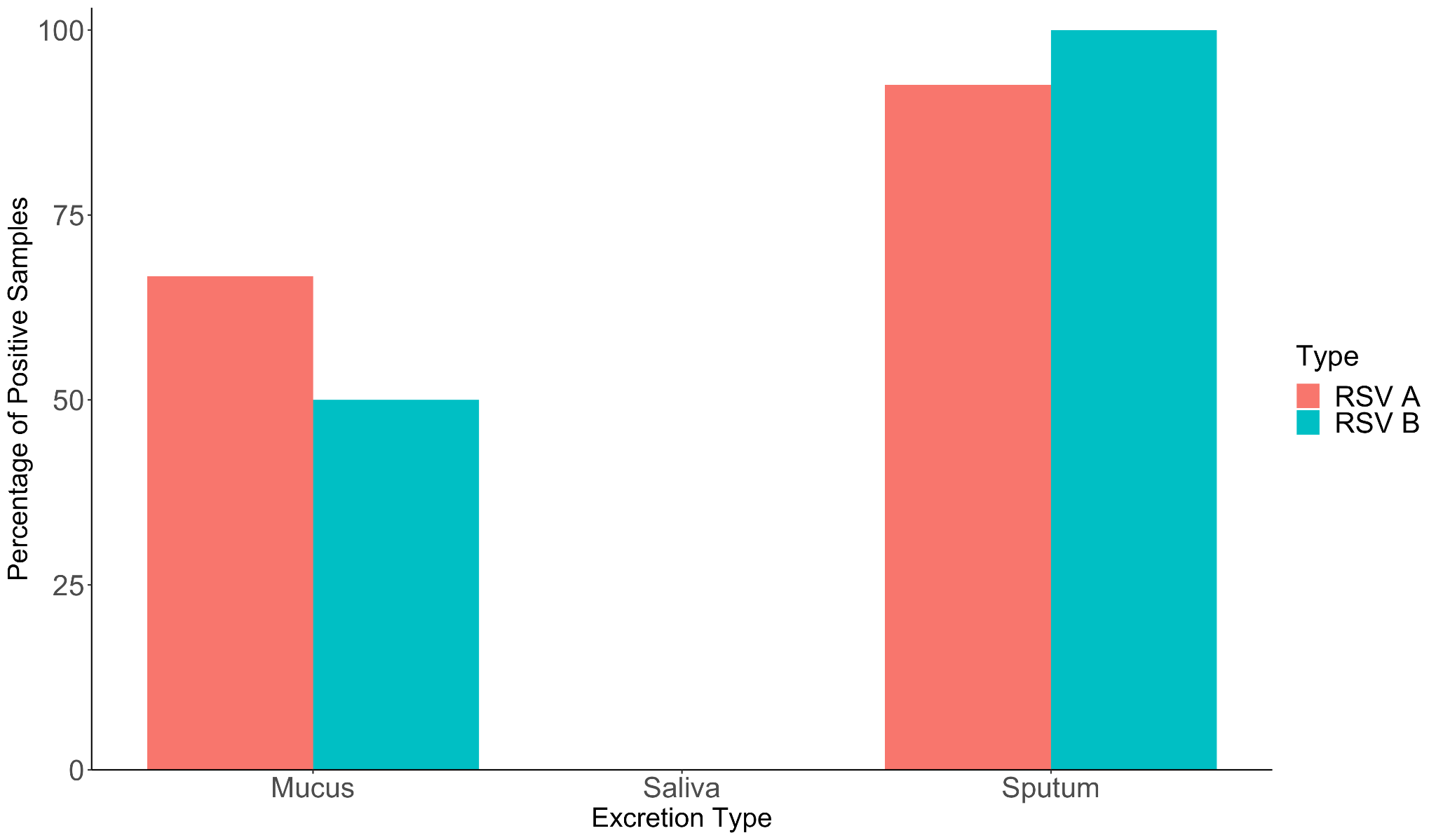

Figure S3. Weighted Averages of Percentage of Positive RSV A or B Detections Separated by RSV Type in Various Excretions from Subjects with Confirmed Infection. Data sets that specified the measurement of one parainfluenza virus type were included (19,20,38); data sets that did not specify the measurement of one RSV type were excluded. Breakdowns for n are as follows: Mucus: A(n=3), B(n=2), Saliva: A(n=0), B(n=1) Sputum: A(n=27), B(n=2). Graphics were created using the statistical computing program R (27).

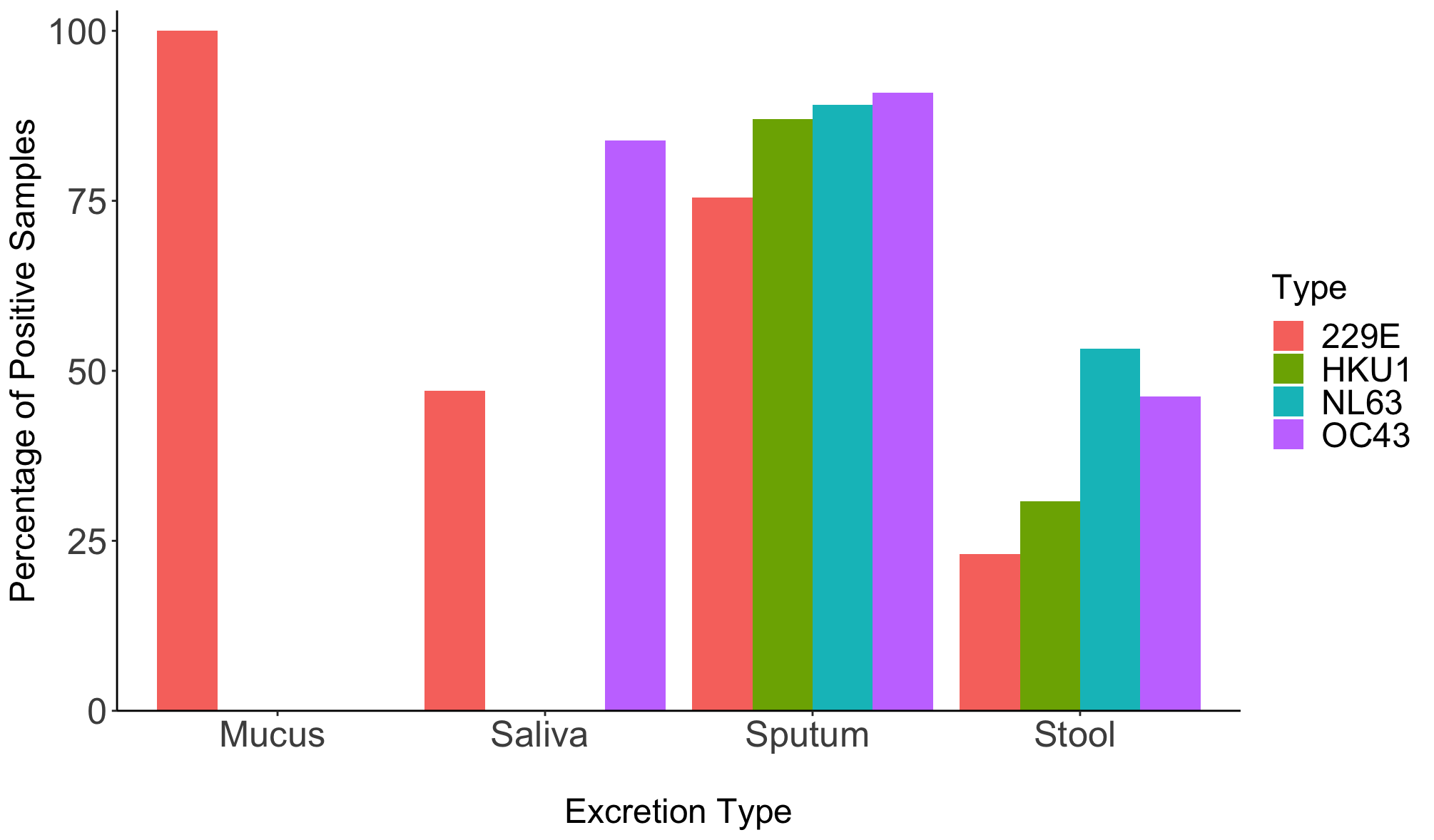

Figure S4. Weighted Averages for Percentage of Positive Seasonal Coronavirus Detections in Various Excretions from Subjects with Confirmed Infection Separated by Seasonal Coronavirus Type. Data sets that specified the measurement of one parainfluenza virus type were included (14,16–20,24,25,31,34,38,50–52); data sets that did not specify the measurement of one seasonal coronavirus type were excluded. Breakdowns for n are as follows: Mucus: 229E(n=1), HKU1(n=0), NL63(n=2), OC43(n=1), Saliva: 229E(n=17), HKU1(n=0), NL63(n=0), OC43(n=56), Sputum: 229E(n=53), HKU1(n=69), NL63(n=92), OC43(n=175), Stool: 229E(n=13), HKU1(n=13), NL63(n=15), OC43(n=13). Graphics were created using the statistical computing program R (27).

28.	Al Khatib H, Coyle P, Al Maslamani M, Al Thani A, Pathan S, Yassine H. Molecular and biological characterization of influenza A viruses isolated from human fecal samples. Infect Genet Evol. 2021;93.

43.	Wang L, Yang S, Yan X, Liu T, Feng Z, Li G. Comparing the yield of oropharyngeal swabs and sputum for detection of 11 common pathogens in hospitalized children with lower respiratory tract infection. Virol J. 2019;16.
